# Learning from the COVID-19 pandemic: a systematic review of mathematical vaccine prioritization models

**DOI:** 10.1101/2024.03.04.24303726

**Authors:** Gilberto Gonzalez-Parra, Md Shahriar Mahmud, Claus Kadelka

**Affiliations:** Instituto de Matemática Multidisciplinar, Universitat Politècnica de València, València, Spain; Department of Mathematics, New Mexico Tech, 801 Leroy Place, Socorro, 87801, NM, USA; Department of Mathematics, Iowa State University, 411 Morrill Rd, Ames, 50011, IA, USA

**Keywords:** Review, mathematical model, age, COVID-19, vaccine allocation, vaccine roll-out

## Abstract

As the world becomes ever more connected, the chance of pandemics increases as well. The recent COVID-19 pandemic and the concurrent global mass vaccine roll-out provides an ideal setting to learn from and refine our understanding of infectious disease models for better future preparedness. In this review, we systematically analyze and categorize mathematical models that have been developed to design optimal vaccine prioritization strategies of an initially limited vaccine. As older individuals are disproportionately affected by COVID-19, the focus is on models that take age explicitly into account. The lower mobility and activity level of older individuals gives rise to non-trivial trade-offs. Secondary research questions concern the optimal time interval between vaccine doses and spatial vaccine distribution. This review showcases the effect of various modeling assumptions on model outcomes. A solid understanding of these relationships yields better infectious disease models and thus public health decisions during the next pandemic.

## 1. Introduction

In December 31, 2019, the World Health Organization (WHO) was informed of several cases of a pneumonia of unknown cause occurring in Wuhan, China (Centers for Disease Control (CDC)). Only 71 days later, the WHO declared after 118,000 cases in 114 countries and 4,291 deaths COVID-19, caused by the Severe Acute Respiratory Syndrome Coronavirus (SARS-CoV2), a pandemic. Despite the widespread implementation of numerous types of non-pharmaceutical interventions (NPIs), aimed at curbing virus spread and keeping hospitals functional, COVID-19 continued to spread rapidly in the absence of a vaccine (Odusanya et al., 2020). Recent advances in mRNA vaccine technology enabled rapid development of highly effective vaccines (Zhang et al., 2019). Since the globalized world had never experienced a pandemic, nor a mass vaccine roll-out at this scale, various formerly mainly theoretical questions related to vaccine access and prioritization became all of a sudden very important. These included: Who should be vaccinated first? Should the second vaccine dose be delayed in order to provide a first dose to more people? What parameters must be taken into account to accurately determine the best prioritization strategy? Should high-income countries share some of their limited vaccine with poorer countries? For ethical reasons only, or does a more equitable vaccine coverage have even epidemiological benefits? In response to these quickly emerging questions, scientists from many fields started to collaborate and suggest answers.

Globally, by the end of 2023, there have been over 770 million confirmed COVID-19 cases and over 7 million deaths, reported to the WHO (World Health Organization (WHO)). Despite the development of highly effective vaccines and 13.6 billion administered COVID-19 vaccine doses, with over 72 percent of the world population having received at least one dose (New York Times), the disease still surges in waves around the world in early 2024. While the infection fatality rate is now substantially lower than in the beginning of the pandemic (Sorensen et al., 2022) and most NPIs have disappeared, large COVID-19 outbreaks and community spread still appear around the world, causing, for example, numerous individuals to suffer from so-called long COVID symptoms that can linger for years post infection (Sudre et al., 2021). Reasons COVID-19 has not disappeared after sufficient production of vaccines include the ongoing emergence of SARS-CoV-2 variants, partial vaccine escape by some variants (Chakraborty et al., 2022; Wang et al., 2021), issues related to vaccine access and distribution specifically in low-income countries (Sheikh et al., 2021), as well as vaccine hesitancy and wariness driven by rampant misinformation (Sallam, 2021). Learning from mistakes made during the COVID-19 pandemic and the first global mass vaccine rollout is thus paramount for future pandemic preparedness.

This review identifies and analyzes a variety of studies related to finding the optimal vaccine allocation given a limited supply. While the specific research questions and settings differ from study to study, age is a crucial factor in all COVID-19 vaccine prioritizations as older people have a substantially higher COVID-19 fatality rate. The primary focus in this review is therefore on studies that were based on a mathematical model, which takes age into consideration. Other important attributes which were used by public health decision-makers to differentiate COVID-19 vaccine access and which are investigated in some of the studies include, among others, occupation (e.g., prioritizing healthcare and essential workers) and comorbidity status (e.g., prioritizing individuals with known risk factors). Moreover, the recommended two-dose vaccine regimen for most COVID-19 vaccines raised the related prioritization question whether it is beneficial to delay the second dose in order to increase initial vaccine coverage. Another related prioritization question concerns spatial aspects (e.g., the optimal distribution of limited vaccine supply between different states or countries). We summarize innovative and interesting mathematical model-based studies that investigate these related prioritization questions, no matter whether the models specifically consider age.

Given the large number of studies related to optimal COVID-19 vaccine allocations, we decided to restrict ourselves to studies that employ a mathematical model for decision-making. The included studies employ several modeling frameworks. Most studies are based on an ordinary differential equation (ODE) model, in which the population is stratified into different compartments. The simplest model, colloquially known as SIR model and first studied nearly 100 years ago (Kermack and McKendrick, 1927), contains three compartments: susceptible (S), infected (I), and recovered (R). More complex models possess additional compartments for individuals that are e.g. infected but not yet infectious, asymptomatically versus symptomatically infected, quarantined but not yet recovered, or dead. To account for different ages and possibly other attributes (e.g., occupation), the population is stratified into a finite number of sub-populations (e.g., age classes) and the compartments are duplicated for each sub-population. Each sub-population can have its own characteristics. This enables modelers to account for heterogeneity (e.g., age dependency) in contact patterns, NPI adherence, vaccine hesitancy, susceptibility to infection, as well as various factors related to disease progression. ODE-based models implicitly make a number of assumptions that are inaccurate for COVID-19 disease dynamics and hard to overcome within the ODE modeling framework. Among others, they assume that (i) the population is homogeneously mixed, (ii) the time spent in each transient compartment is exponentially distributed, and (iii) disease dynamics are deterministic.

Another modeling framework, agent-based models (ABMs; also known as individual-based models), is stochastic in nature and employed by a smaller number of studies. In ABMs, individual agents (i.e., people) are modeled; agents interact with each other and possibly spread the disease through e.g. heterogeneous interaction networks. This modeling framework is highly flexible (e.g., each individual can have its own characteristics and decision rules) and can be adaptive (e.g., the decisions of an agent can depend on other’s decisions). However, ABMs inherently rely on simulations. Their stochastic nature further increases the computational needs, rendering an exhaustive exploration of a large parameter space impossible. Lastly, a few studies employ partial differential equation (PDE) models. These studies typically focus on spatial aspects of vaccine prioritization.

Contrary to other review articles on this topic (Saadi et al., 2021; Liu and Lou, 2022; Noh et al., 2021; Thakkar and Spinardi, 2023), the focus of this review is on understanding the effect of modeling assumptions and parameters on policy recommendations. For example, while most studies agree that elderly and vulnerable should be vaccinated first due to their substantially higher infection fatality ratio, some studies suggest the opposite. We look in detail at which model parameters and assumptions cause these discrepancies. This review includes 94 articles, which use a mathematical model to answer at least one of three questions related to COVID-19 vaccine prioritization:

1. How should a limited vaccine be optimally distributed among a population stratified by age (and possibly other factors)?
2. For limited vaccines with a two-dose regimen, should the second dose be delayed in order to provide more people with a first vaccine dose?
3. How should a limited vaccine be optimally distributed given spatial heterogeneity?

In Section 2, we briefly describe how we identified articles of interest. Section 3 summarizes, at a high-level, the main findings of these articles related to vaccine prioritization. Section 4 puts these findings into context, particularly those without a clear consensus strategy. Several key COVID-19 model parameters and assumptions are introduced, with a focus on how they affect optimal vaccine prioritization strategies. Section 5 provides a brief summary of particularly interesting and noteworthy studies. Finally, Section 6 briefly presents related works that employ optimal control methods to answer questions related to vaccine prioritization.

## 2. Methods

To find studies of interest, we searched PubMed, the Web of Science Core Collection and MathSciNet (all in February 2024) for research articles that contain the following keywords: ‘age’ AND ‘model’ AND (’COVID-19’ OR ‘SARS-CoV-2’) AND (’vaccine’ OR ‘vaccination’) AND (’best’ OR ‘optimal’ OR ‘priorit*’) AND (’mathematical’ OR ‘computational’ OR ‘stochastic’ OR ‘network’). After removing duplicates (e.g., preprints and journal articles) and non-peer-reviewed preprints, this yielded a total of 285 articles, which we manually reviewed, in addition to 43 articles known to the authors and/or referenced in one of the 285 articles (Fig. 1A). For each article, we decided if it contained a mathematical model that answers at least one of the COVID19 vaccine prioritization questions stated above. This yielded a total of 94 articles included in this review (Fig. 1B). Any article that did not assume limited vaccine availability (e.g., studies looking into the epidemiological effect of boosters in high-income countries) was excluded.

**Figure 1:**
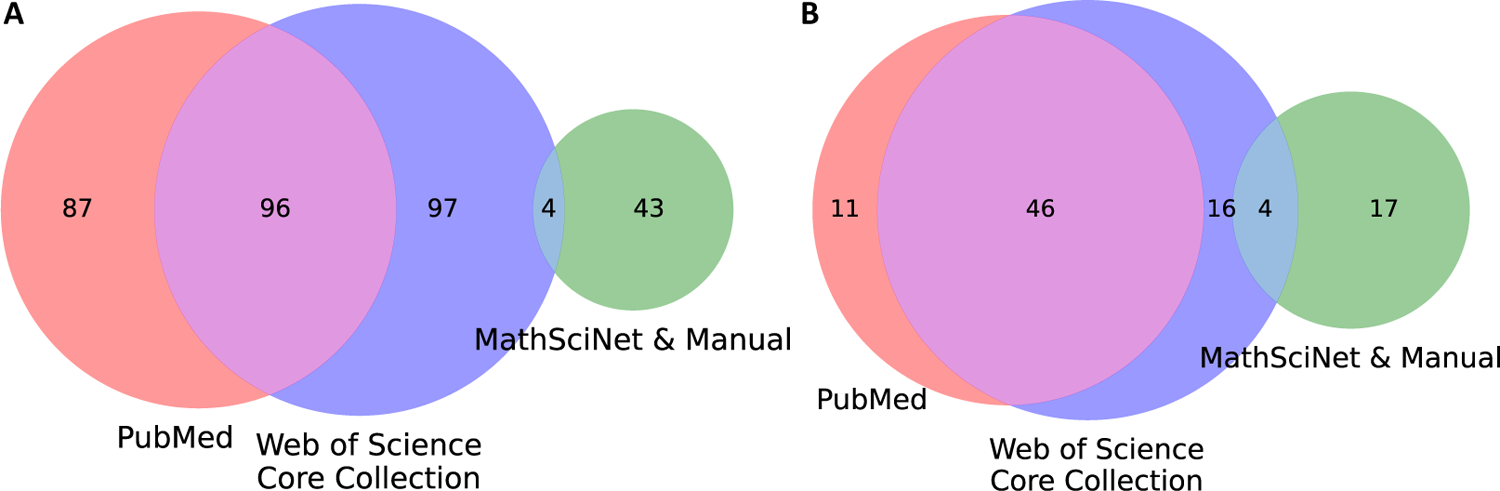
Number of (A) investigated and (B) included studies, stratified by source.

Eighty of the included articles contain an age-stratified mathematical model that provides answers to our primary research question: How should a limited vaccine be optimally distributed among a population stratified by age? Fifteen articles contain a mathematical model (not neccessarily agestratified) to answer the secondary research question: For limited vaccines with a two-dose regimen, should the second dose be delayed in order to provide more people with a first vaccine dose? Finally, seven articles contain a mathematical model that considers spatial aspects of vaccine distribution. The low number of articles related to the latter two research questions is, at least partially, due to the fact that the keywords were selected to preferentially find articles investigating our primary research question.

## 3. Summary of findings

Collectively, the included articles contain models that are tailored to cities, states or countries from all continents except Antarctica (Fig. 2). A few studies tailor their model to more than one country to showcase how variability in e.g. age distributions, age-stratified contact patterns, or implemented NPIs can affect optimal vaccine prioritization, see e.g., Gozzi et al. (2021); Liu et al. (2022b); Wang et al. (2022); Liu et al. (2022a). Other studies employ more abstract models, frequently ABMs, which are not tailored to any specific setting, see e.g., Romero-Brufau et al. (2021); Grauer et al. (2020); Kadelka and McCombs (2021). These models contain tuneable parameters and are well-suited to reveal the qualitative dependence of optimal allocation strategies on key parameters and assumptions.

**Figure 2:**
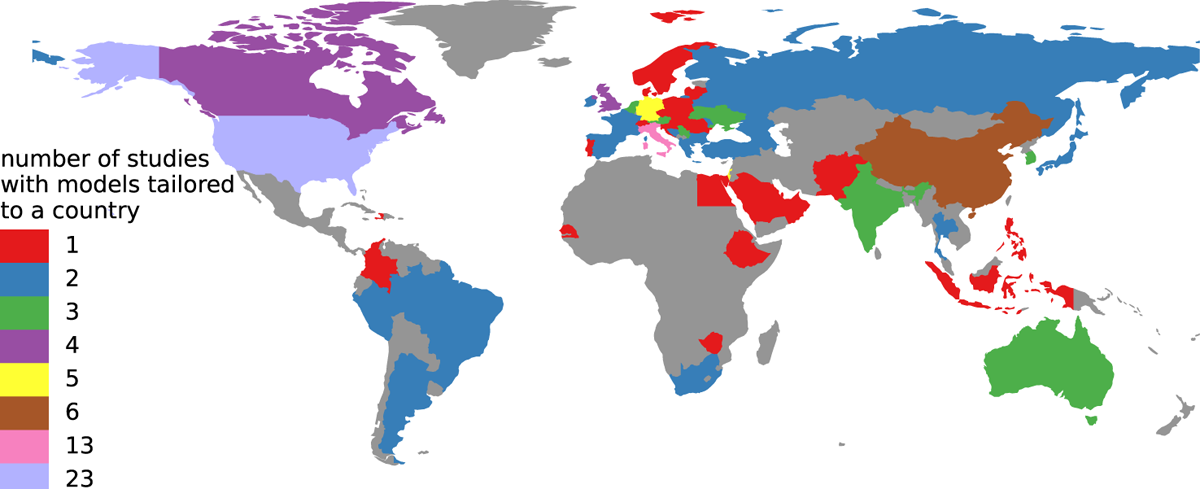
Number of studies that contain a model for a specific country. Some studies include models tailored to several countries, while others are more abstract and not tailored to a specific country. Census data and age-stratified contact matrices are two examples of frequently used country-specific data.

While the included articles employ a variety of metrics to quantify the quality of a given vaccine allocation strategy, there are several common objectives. In decreasing order of use (Fig. 3), these include: minimizing deaths (used in 80 of the 94 included studies), cases/infections (56), hospitalizations (22), and years of life lost (YLL; 10). Other, less frequently used objectives include minimizing qualityand disability-adjusted life years (QALYS and DALYS, respectively; used in 6 studies), minimizing the peak number of hospitalized, as well as several equitable and economic considerations. While technically different and considered as separate objectives in at least one study (Islam et al., 2021), we do not differentiate between the objectives minimizing cases and infections. Some studies attempt, furthermore, to optimize multiple objectives at the same time, e.g., through the use of optimal control methods (summarized in Section 6), or an analysis of Pareto-optimal allocation strategies.

**Figure 3:**
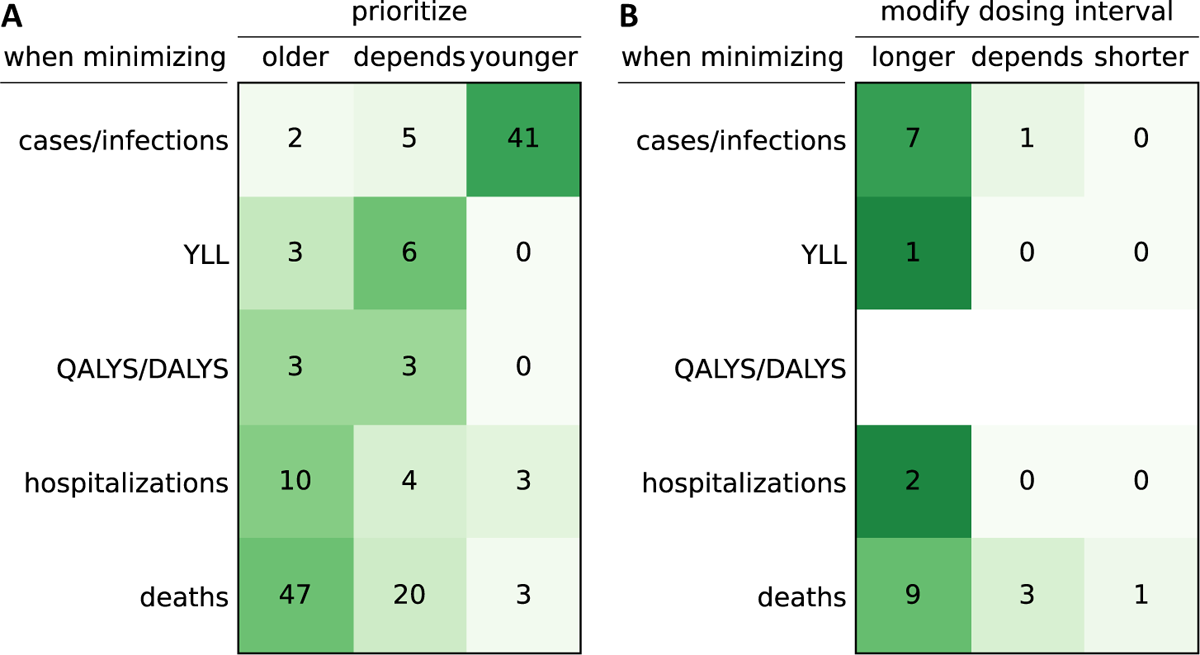
High-level summary of findings. **(A)** Number of studies that agree at a high-level on a given prioritization strategy (columns) when minimizing a given metric (rows). Only studies that are based on a mathematical model that considers stratifying vaccine access by age are included. Note that all studies that recommend prioritization of the oldest and most vulnerable people, possibly after vaccinating health care workers, were nevertheless counted as prioritizing older. **(B)** Number of studies that agree at a high-level on a dosing interval strategy (columns) when minimizing a given metric (rows). (A-B) The second column (“depends”) includes all studies that present more subtle findings where the prioritization and dosing interval depends on certain assumptions. In Table 1 and Table 2, the high-level summaries are stratified by study.

**Table 1:**
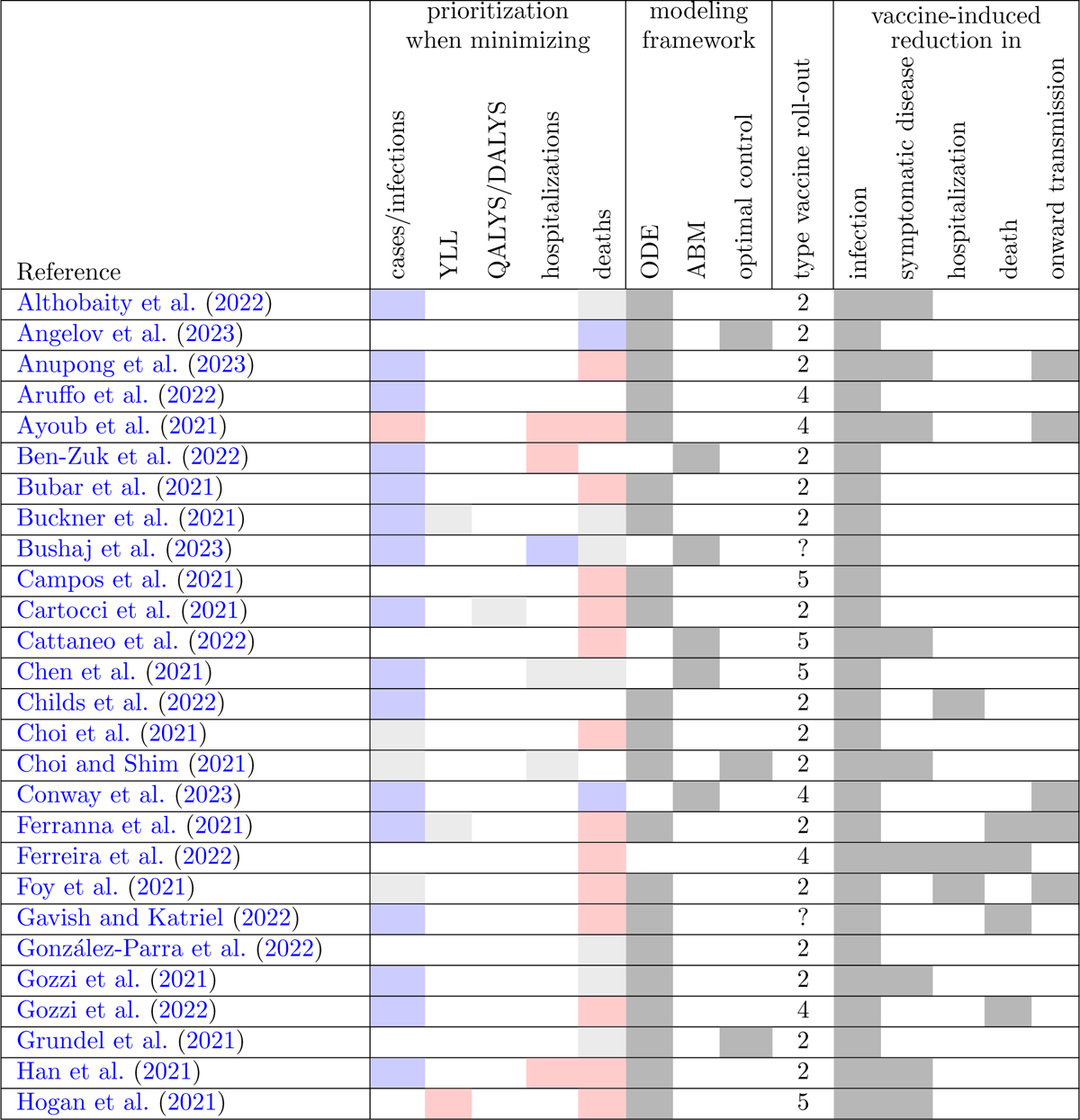
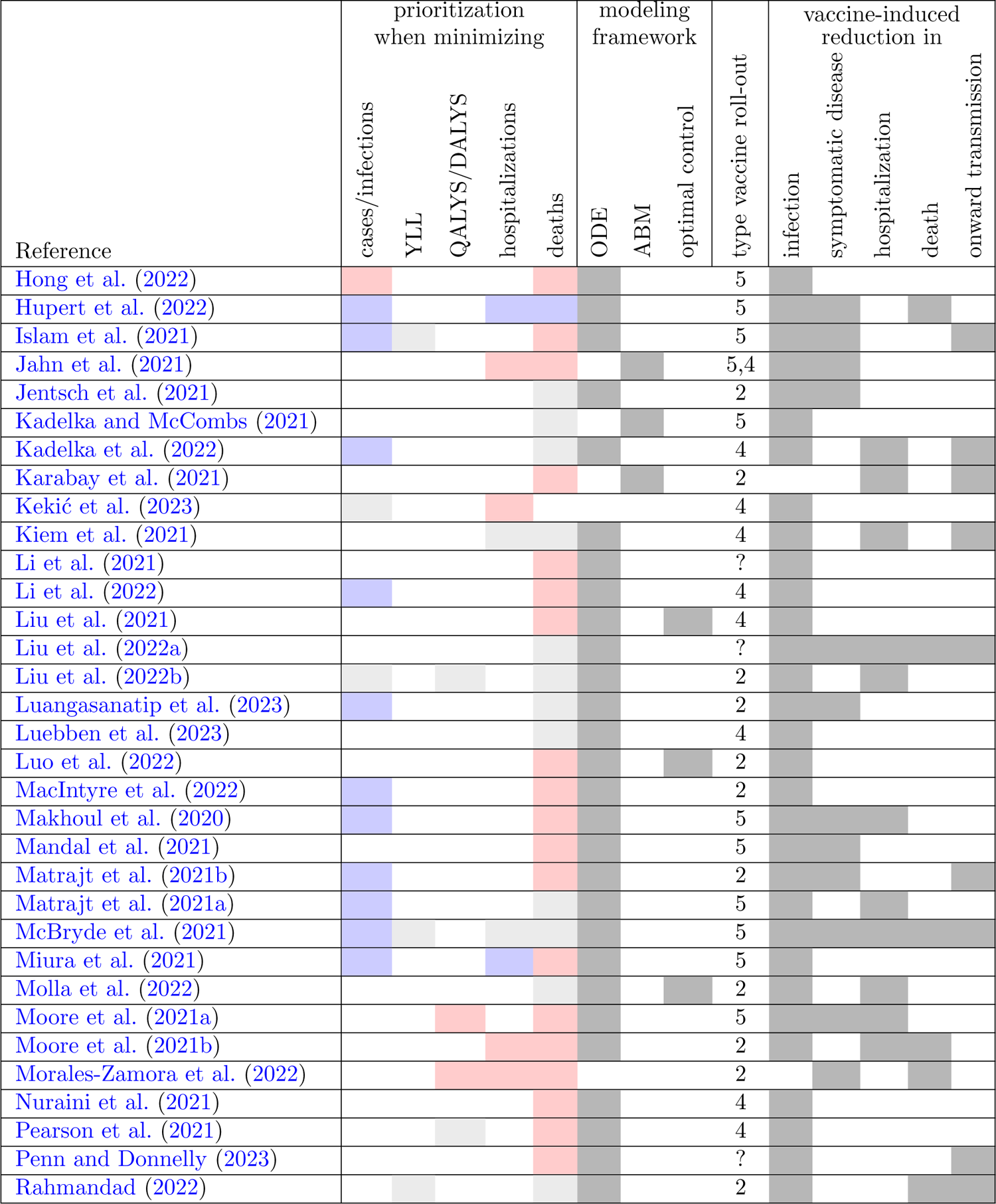
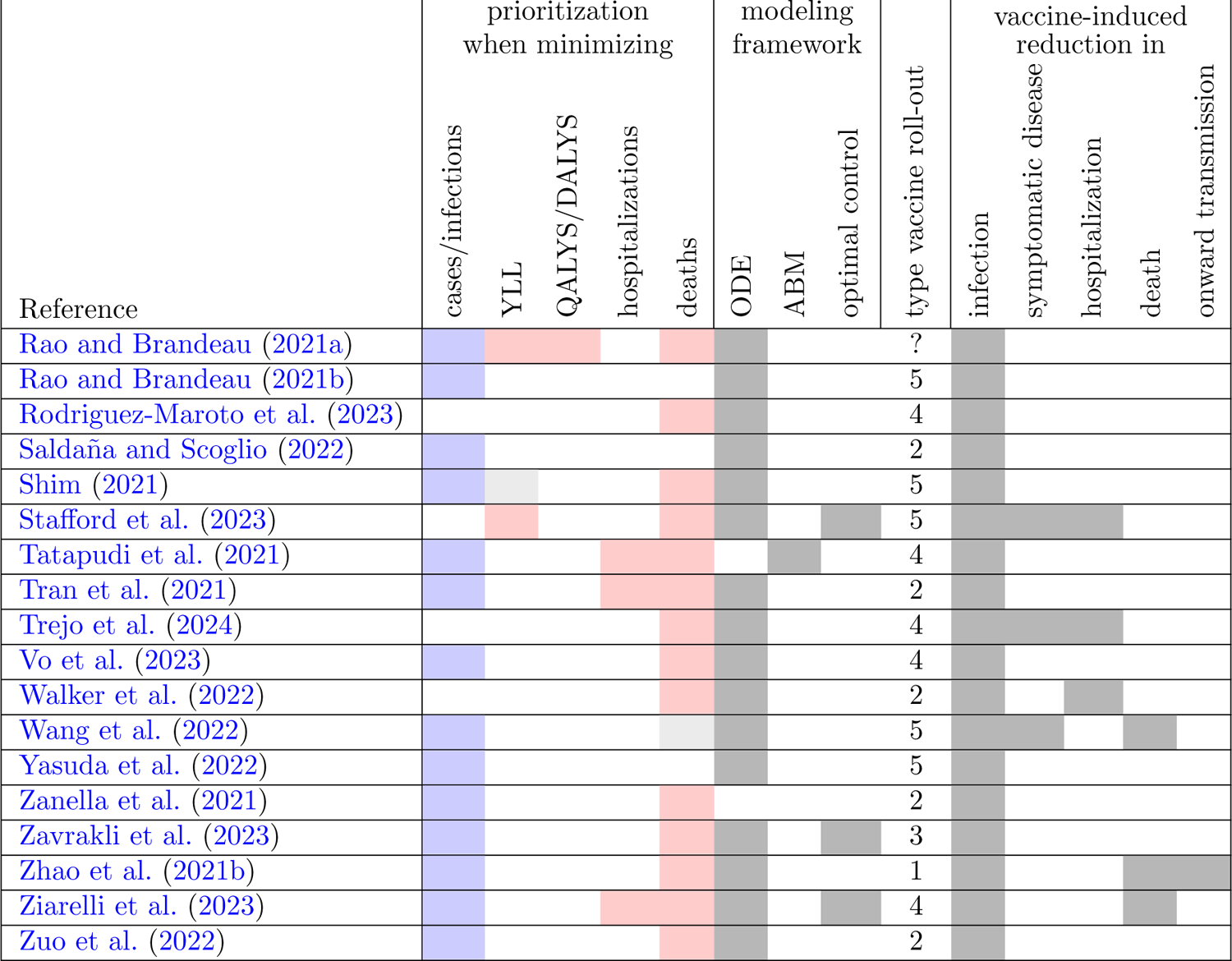
High-level summary of COVID-19 vaccine prioritization studies. Each row summarizes the high-level prioritization strategy identified by a given study (white: not assessed, red: prioritize older/vulnerable population, blue: prioritize younger/highcontact population, gray: prioritization depends on model assumptions). Fig. 3A provides summary counts. Only studies that were based on a mathematical model that accounts for age were included. The models are further classified by framework, type of vaccine roll-out (as specified in 4.4), as well as considered vaccine functions.

**Table 2:**
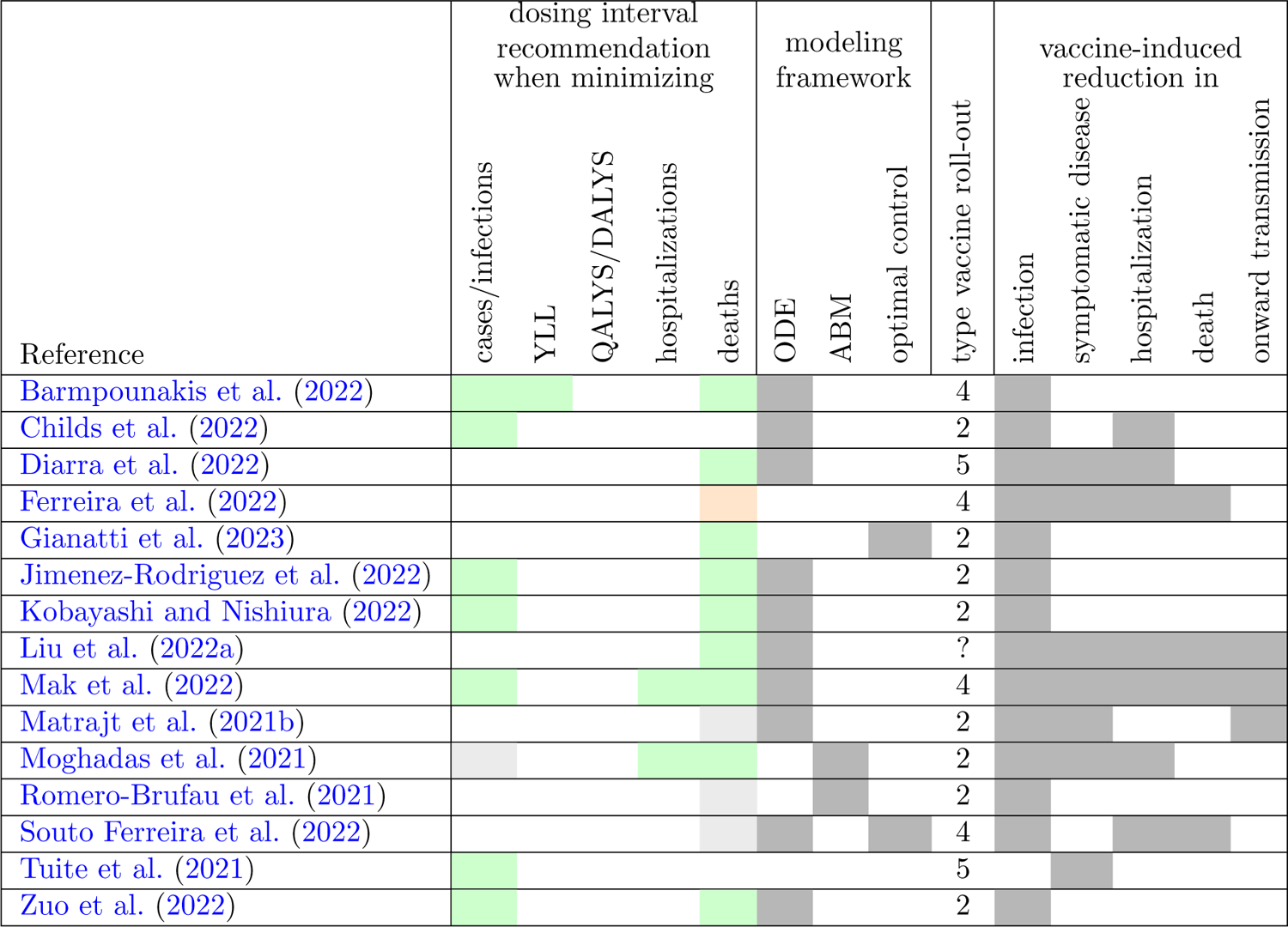
High-level summary of COVID-19 vaccine dosing interval studies. Each row summarizes the high-level dosing interval recommendation identified by a given study (indexed by reference number). White: not assessed, green: delay second dose, orange: shorten dosing interval, gray: recommendation depends on model assumptions. Fig. 3B provides summary counts. Only studies that were based on a mathematical model were included. The models are further classified by framework, type of vaccine roll-out (as specified in 4.4), as well as considered vaccine functions.

When minimizing mortality is the sole objective, the majority of studies (47 out of 70) agree that vaccinating older individuals, vulnerable individuals, and if considered health care workers first is optimal, irrespective of the specific setting or assumptions (Fig. 3A, Table 1). There exists, however, some disagreement about the prioritization among these subpopulations. Interestingly, 23 model-based studies (32.9%) conclude that under certain circumstances a prioritization of younger people who have on average more contacts leads to lower death counts. Qualitatively, the optimal prioritization strategies do generally not shift much when minimizing other morbidity-based metrics such as YLL or hospitalizations. On the other hand, most studies (41 out of 48, 85.4%) agree that to minimize the total number of infections and/or the effective reproductive number younger individuals should be vaccinated first since they typically have more contacts and thus more chances to spread the virus (Fig. 3A).

The recommended dosage for some of the most effective and initially most widely available COVID-19 vaccines, e.g., the Pfizer-BioNTech, the Moderna, and the AstraZeneca vaccine, was two doses. While a single dose offers some protection, two doses, spaced out at least a few weeks, induce a substantially stronger protection. Thus, a related prioritization question concerns the optimal allocation of each individual vaccine dose. If the vaccine supply is limited, a delay of the second dose allows for more individuals to receive a first dose. A total of 15 studies (not necessarily age-structured) investigated this particular prioritization question. Most studies agree that a delay of the second dose is beneficial, irrespective of the specific objective (Fig. 3B, Table 2). This aligns with findings from a pooled analysis of four randomised trials (Voysey et al., 2021). Several studies identify the relative protection induced by the first dose compared to the full vaccine regimen as a key parameter in this decision (Romero-Brufau et al., 2021; Matrajt et al., 2021b; Souto Ferreira et al., 2022), highlighting the need for detailed vaccine effectiveness data.

Countries are spatially heterogeneous. Thus, spatial factors can affect the optimal allocation of limited vaccine. While not the primary objective of this review, we identified a number of studies that investigate spatial aspects of vaccine distribution (Table 3). The considered questions are more diverse than in the previous two research questions; we therefore provide most details in Subsection 5.3. In summary, the investigated studies all agree that spatial factors are important when designing deaths-minimizing optimal vaccine prioritization plans and that non-trivial trade-offs emerge, e.g. between prioritizing regions with high incidence counts whose inhabitants are on average younger and regions with more retirees. Economic factors are also taken into consideration by multiple studies.

**Table 3:**
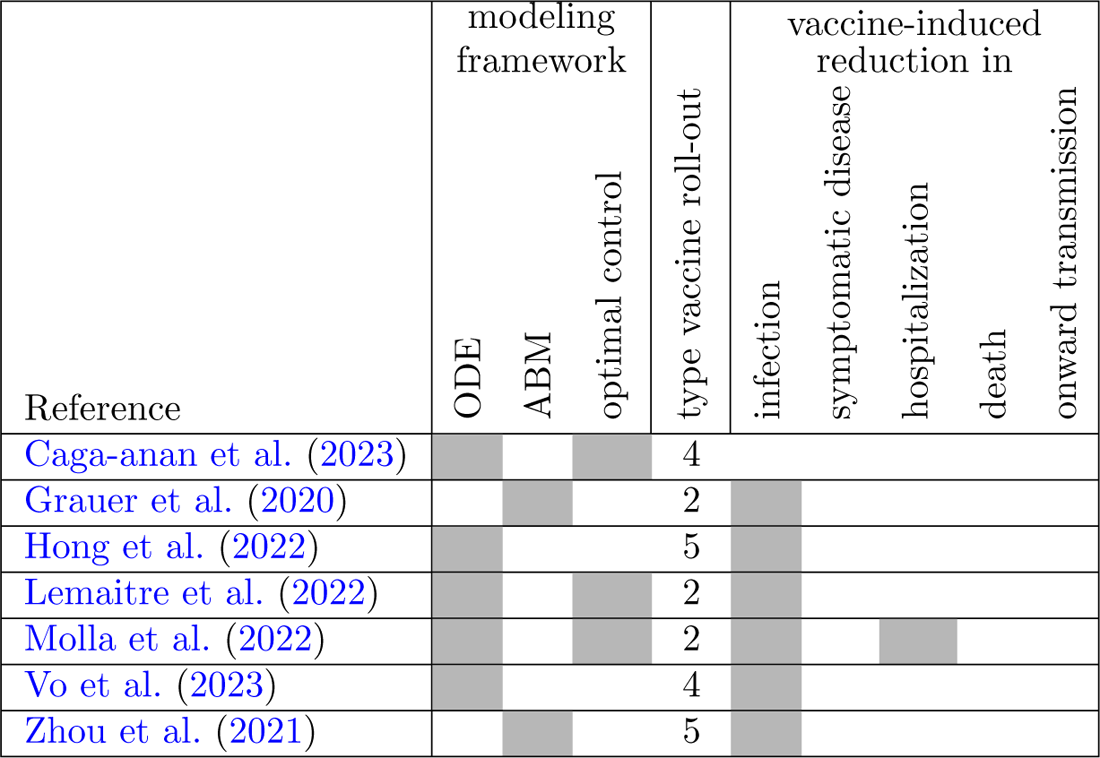
Summary of spatial COVID-19 vaccine distribution studies. Each row describes a study that developed a spatial vaccine prioritization model. The models are classified by framework, type of vaccine roll-out (as specified in 4.4), as well as considered vaccine functions.

## 4. Key implementation details in vaccine prioritization models

Modelers make many decisions some consciously, some unconsciously when creating a mathematical vaccine prioritization model. Some choices can fundamentally affect the resulting optimal vaccine allocation. In this section, we focus on the studies that identify a dependence of the optimal prioritization strategy (Fig. 3) to better understand the effect of certain modeling assumptions as well as the impact of setting-to-setting differences in key parameters.

### 4.1. Modeling framework

The choice of modeling framework may affect outcomes of vaccine prioritization models. In particular, as briefly described in the introduction, the popular ODE-based compartmental models come with several implicit assumptions. The homogeneous mixing assumption can be overcome by stratifying the population into sub-populations and accurately describing heterogeneous mixing (see Subsection 4.6). Another implicit assumption of ODE-based models is that the time spent in each transient compartment is exponentially distributed. This is frequently unrealistic. For example, upon infection with SARS-CoV-2 the virus needs time to replicate before a person becomes contagious. The latent period is therefore not exponentially distributed Zhao et al. (2021a). While most included studies ignore this issue likely since there exists no apparent direct effect on vaccine prioritizations -, some studies stratify a single transient compartment into multiple (see e.g., Moore et al. (2021b); Childs et al. (2022)). This has the effect that the total time spent in these compartments follows an Erlang distribution (assuming equal average time in each of the multiple compartments) rather than an exponential distribution. The Erlang distribution, as a special case of the Gamma distribution, is more flexible and can thus describe more accurately the average time an individual is e.g. latently infected with SARS-CoV-2 (Lloyd, 2001).

### 4.2. Prediction horizon

Public health decision makers typically operate within a defined planning horizon. That is, they attempt to make decisions that yield “optimal” outcomes over the course of a given time period. Similarly, mathematical models compare outcomes (e.g., total deaths or cases under different vaccine allocation strategies) over a defined time interval whose length is known as prediction horizon. The main benefit of a short prediction horizon is reduced uncertainty since long-term disease dynamics are very difficult to predict. A German ODE-based study nicely highlights that the choice of prediction horizon fundamentally influences who to vaccinate first (Grundel et al., 2021). If the horizon is too short (less than 8 weeks in the study), prioritization targets may switch as the strategy suffers from shortsightedness. Another study shows that vaccinating elderly is always preferred for a short prediction horizon, which may however yield sub-optimal long-term outcomes (Campos et al., 2021).

### 4.3. Vaccine eligibility

Given an initially limited COVID-19 vaccine supply, people with a known history of COVID-19 infection were excluded from early vaccine access by most public health agencies. Correspondingly, most reviewed mathematical models assume that only susceptible individuals can be initially vaccinated. Some ODE-based studies with more compartments (see e.g., Islam et al. (2021); Karabay et al. (2021); Taboe et al. (2023); Luo et al. (2022); Anupong et al. (2023); Grundel et al. (2021)) allow for vaccination of any individuals without known COVID-19 history. That means preor asymptomatically infected as well as recovered individuals without known history of infections (e.g., through positive test results or symptoms) are also eligible for early vaccination, leading to some vaccine doses being used sub-optimally. A few studies even quantify the reduction in deaths, YLL, and infections that could be achieved through the hypothetical use of seroprevalence tests prior to vaccination (Bubar et al., 2021; Ayoub et al., 2021). While challenging to implement, these studies find, as expected, that vaccinating only seronegative individuals always leads to improved outcomes, with the difference being larger at higher levels of seroprevalence.

### 4.4. Vaccine roll-out

When deciding who to vaccine first, public health officials must anticipate the speed of the vaccine roll-out. In mathematical models, this results in assumptions about the daily number of vaccinations. Post-hoc analyses benefit from access to historic vaccination data and can simply ask the question: Given this number of vaccinations per day, how could these vaccines have been allocated in an optimal way? When such data is unavailable, e.g., prior to the start of a mass vaccine roll-out, modelers typically make one of the following assumptions in ODE models. Here, let *X* = *X*(*t*) denote the subset of the population that is eligible for vaccination (e.g., all susceptibles) at time *t*. Then, vaccination of part of these eligible individuals can be described by

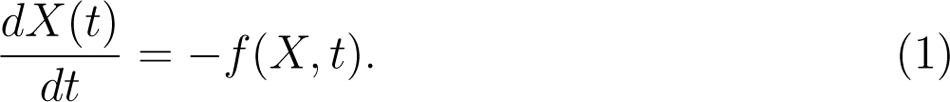

Note that this equation only considers the vaccination process. The size of *X* may also change due to natural infection, immunity waning, etc. The rate of newly vaccinated, *f* (*X, t*), typically takes one of the following forms:

1. *f* (*X, t*) = *νX*(*t*), where *ν ≥* 0 describes the proportion of eligible individuals vaccinated per unit time. This form implies that the number of vaccinations is proportional to the size of *X*. Specifically, as the size of *X* decreases over time (due to vaccination, natural infection, etc.), the number of newly vaccinated decreases as well. Mathematically, this form guarantees that *X*(*t*) remains positive for all time. This form is used in Zhao et al. (2021b); Acuña-Zegarra et al. (2021).
2. *f* (*X, t*) = *c,* where *c ≥* 0 is a constant that describes the number of vaccinations per unit time. Mathematically, this form does not guarantee positivity of *X*(*t*) for all time, as all individuals may eventually become vaccinated (or otherwise removed from *X*). This necessitates careful attention when numerically solving the ODE. Nevertheless, this form is used in many models (Table 1), likely due to its simplicity.
3. *f* (*X, t*) = *ν*(*t*)*X*(*t*). This form is the most complex. The proportion of eligible individuals being vaccinated may vary over time. This form has the same nice mathematical property as form 1: positivity of *X*(*t*) is guaranteed for all time. Contrary to form 1 and form 2, this more complex third form allows for the rate and also the number of vaccinations to increase over time, as is typically the case at the beginning of a mass vaccine roll-out. Form 1 (and form 2), on the other hand, assume that the number of vaccinations decreases (remains constant, respectively) as the number of eligible individuals decreases. This form is used in a few models that employ optimal control techniques (Acuña-Zegarra et al., 2021; Zavrakli et al., 2023).
4. *f* (*X, t*) = *c*(*t*). In this form, the number of vaccinations only depends on time but not on the size of *X*. This form is well-suited for post-hoc analyses, in which the number of vaccinations that were conducted per unit time (e.g., day or week) is known, see e.g. Islam et al. (2021); Gozzi et al. (2022); Luebben et al. (2023); Kekíc et al. (2023); Aruffo et al. (2022); Ziarelli et al. (2023); Ferreira et al. (2022); Cattaneo et al. (2022). Mathematically, this form requires careful attention when solving the ODE numerically, to ensure *X*(*t*) remains non-negative at all time. This can be achieved by adding a number of model constraints, as in Han et al. (2021). One study tailors an ODE model to three different Indonesian provinces and optimizes the function *c*(*t*) such that active cases remain below an acceptable threshold and total vaccination cost is minimized; interestingly, the optimal function *c*(*t*) is highly nonmonotonic (Nuraini et al., 2021). Another study optimizes *c*(*t*) as well, by assuming that vaccines are produced at a constant speed but that vaccine stock needs not to be used immediately (Souto Ferreira et al., 2022).
5. A number of studies do not specify *f* (*X, t*). Rather, they assume that all vaccinations have been completed prior to the simulation of the disease spread. This simplifying assumption decouples the vaccine rollout from the disease spread. A modified version of this approach is implemented in Matrajt et al. (2021b) where the simulation of disease dynamics is stopped once a week when a specified number of (weekly) vaccinations occur. A similar approach is implemented in an ABM in Jahn et al. (2021).

Despite different implementations of the vaccine roll-out, model-based studies generally agree that prioritization of younger, high-contact individuals may be beneficial and even lead to fewer deaths, when the entire vaccine roll-out takes place very quickly, i.e., when vaccines for a large proportion of the population are available quickly (Matrajt et al., 2021a; Buckner et al., 2021; Liu et al., 2022b; McBryde et al., 2021). In this case, the vulnerable population is protected indirectly, by reaching herd immunity and a stop of community spread. This strategy becomes particularly reasonable in situations with low community spread (i.e., the effective reproductive number R_eff_ *≈* 1) (Chen et al., 2021; Althobaity et al., 2022; Gozzi et al., 2021) and in which the epidemic is already in decline (i.e., *d*R_eff_(*t*)*/dt <* 0) (Molla et al., 2022). One prominent study agrees that younger individuals should only be prioritized, when minimizing deaths, if effective reproductive numbers are low but finds that a slow roll-out (and not a fast one) is an additional requirement (Bubar et al., 2021).

### 4.5. Vaccine function and efficacy

In theory, vaccines can improve outcomes in a variety of ways. A vaccinated individual may be less likely (than an unvaccinated individual with same characteristics) to (i) become infected, (ii) experience symptoms when infected, (iii) require hospitalization due to severe symptoms, (iv) die. In addition, a vaccinated person (v) may be less contagious (e.g., due to a lower average viral load), and (vi) may have a shorter duration of infectiousness, i.e., faster disease progression. To illustrate how these different vaccine functions are frequently included in compartmental models, consider the following COVID-19 model, which stratifies the population by disease status (susceptible (S), recently infected but not yet infectious (E=exposed), symptomatically infected (I), asymptomatically infected (A), severely infected/requiring hospitalization (H), deceased from COVID-19 (D), recovered (R)) and vaccine status (superscript v for vaccinated)):

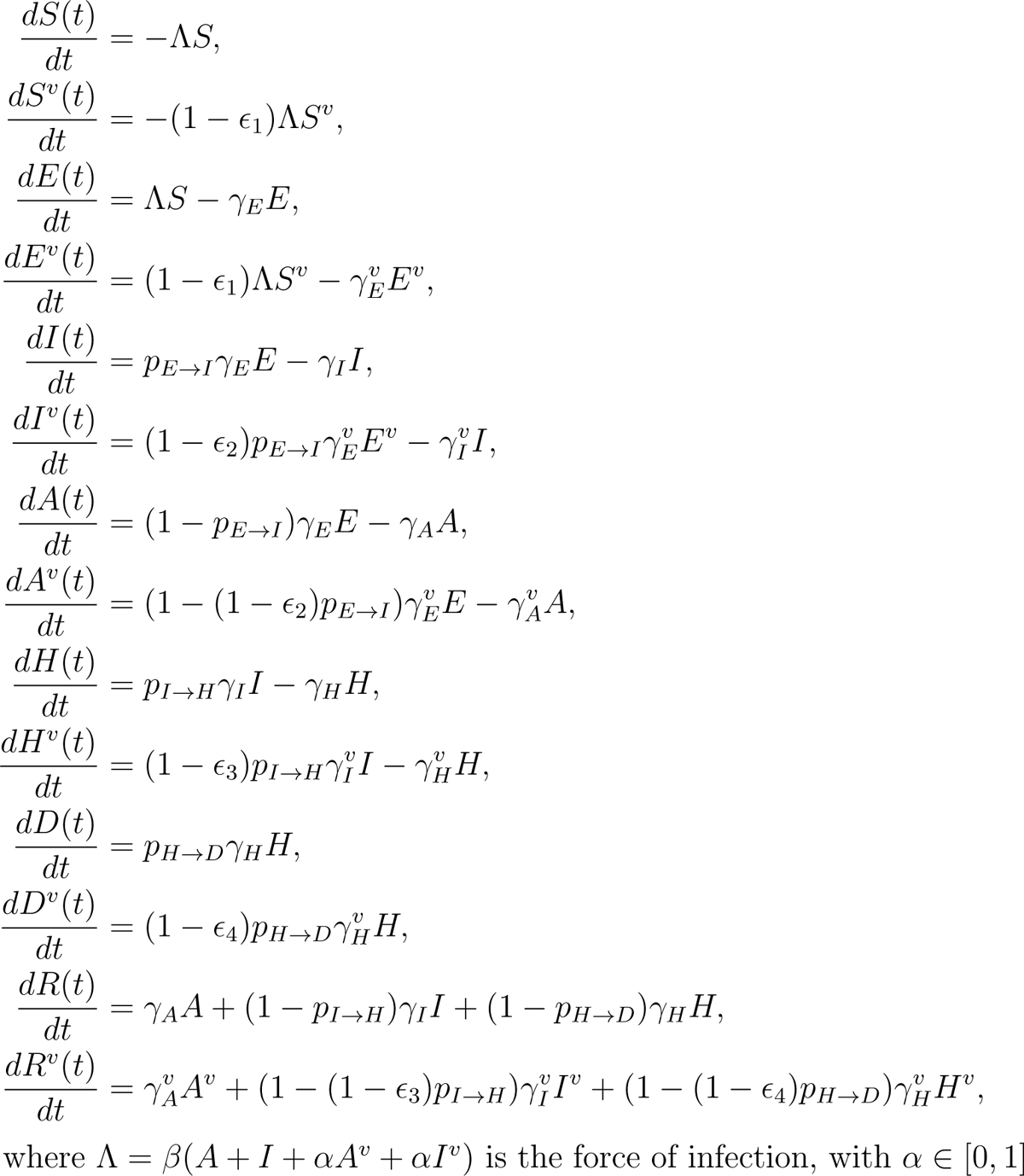

describing the vaccine-induced reduction in onward transmission. Vaccine-induced faster disease progression may be implemented by *γ^v^ ≥ γ_x_* for *x ∈ {E, I, A, H}*.

Three important notes: First, this model implements a so-called leaky vaccine: any vaccinated individual may still become infected, at a lower rate than unvaccinated. A leaky vaccine represents the most frequent implementation of vaccine function. An alternative, also frequently observed assumption is an all-or-nothing vaccine. In that case, *ɛ*_1_ determines the fraction of vaccinated individuals that are completely immune to infection, while the remaining proportion of vaccinated (1 *− ɛ*_1_) are typically assumed to be as susceptible as unvaccinated individuals. In some models, their susceptibility is reduced by a certain degree. Second, a stratification by age can easily be included by duplicating all compartments for each age group, including contact patterns in the force of infection, and considering age-dependent parameters. Third, the parameters *ɛ*_2_*, ɛ*_3_*, ɛ*_4_ describe conditional probabilities. For example, *ɛ*_2_ describes the reduction in symptomatic disease among infected vaccinated compared to infected unvaccinated individuals. The overall vaccine-induced reduction in symptomatic disease, measured in clinical trials and commonly referred to as vaccine efficacy (Halloran et al., 1997), is thus VE_COVID_ = 1 *−* (1 *− ɛ*_1_)(1 *− ɛ*_2_). Similarly, the overall vaccine-induced reduction in deaths is VE_death_ = 1 *−* (1 *− ɛ*_1_)(1 *− ɛ*_2_)(1 *− ɛ*_3_)(1 *− ɛ*_4_).

Used in 90 of the 94 investigated models (Table 1), reduction in infection (*ɛ*_1_, implemented either as a leaky or all-or-nothing vaccine) is the most frequently considered vaccine function, followed by reduction in symptoms (*ɛ*_2_, used in 28 studies), reduction in severe disease (*ɛ*_3_, used in 22 studies), reduction in onward transmission (*α*, used in 17 studies), and reduction in death (*ɛ*_4_, used in 15 studies). Other vaccine functions considered in only a few models include a shorter period of infectiousness (Makhoul et al., 2020; Penn and Donnelly, 2023), as well as a reduced vaccine efficacy for older individuals (Bubar et al., 2021; Aruffo et al., 2022; Buckner et al., 2021) and children (Han et al., 2021). 46 out of 94 studies (48.9%) considered only one type of vaccine function, while three studies (Liu et al., 2022a; McBryde et al., 2021; Mak et al., 2022) differentiated five types (Table 1).

The range of parameter values considered for a given vaccine function also varied wildly. One study investigated optimal prioritization strategies for mass vaccinations with commonly used vaccines that had shown some beneficial heterologous effects against SARS-CoV-2 infection (Hupert et al., 2022). This study considered *ɛ*_1_*, ɛ*_2_*, ɛ*_4_ *∈* [5%, 15%]. In line with results from COVID-19 vaccine clinical trials, most studies assumed relatively high levels of vaccine efficacy against symptomatic disease, VE_COVID_, with the specific values varying based on vaccine product, number of doses and predominant virus variant. Moreover, the relative contribution of *ɛ*_1_ and *ɛ*_2_ differs, with some studies (see e.g., Islam et al. (2021); Matrajt et al. (2021b,a); Han et al. (2021); Ayoub et al. (2021); Makhoul et al. (2020); Choi and Shim (2021); Hogan et al. (2021); Moore et al. (2021a); Liu et al. (2022b); Jahn et al. (2021); Kiem et al. (2021)) contrasting optimal vaccination strategies for both extreme cases: *ɛ*_1_ = VE_COVID_*, ɛ*_2_ = 0 (sterilizing vaccine), and *ɛ*_1_ = 0*, ɛ*_2_ = VE_COVID_ (non-sterilizing vaccine). These studies agree that at a fixed (overall) vaccine efficacy against symptomatic disease, higher *ɛ*_1_ (i.e., lower *ɛ*_2_) leads to better outcomes. The higher *ɛ*_2_ relative to *ɛ*_1_, the more important is the prioritization of older and vulnerable people when optimizing morbidity-based metrics (Islam et al., 2021; Choi and Shim, 2021; Liu et al., 2022b; Kiem et al., 2021). Although hard to disentangle in practice, it is therefore important for optimal prioritization design to understand the relative contribution of *ɛ*_1_ and *ɛ*_2_ to the vaccine efficacies observed in clinical trials.

Studies which differentiate between single-dose and “fully” vaccinated individuals include two parameters for each vaccine function. One study shows that, for a fixed VE_COVID_, delaying second doses and thus covering a larger part of the population with first doses becomes more important at higher *ɛ*_1_ (i.e., lower *ɛ*_2_) when minimizing mortality (Matrajt et al., 2021b). Another important factor is the speed of the vaccine roll-out. One study shows that a generally delayed second dose only leads to fewer deaths if the roll-out is slow (Romero-Brufau et al., 2021). An age-dependent strategy (providing two doses to people 65 and older but delaying the second dose for younger people) performs consistently well, irrespective of the speed of the roll-out. The most important parameter in determining whether a delay of second doses is beneficial is, however, the relative difference in the reduction in susceptibility after one dose versus two doses. As expected, all studies agree that a delay becomes more beneficial the smaller the difference, irrespective of the optimization objective (Romero-Brufau et al., 2021; Moghadas et al., 2021; Mak et al., 2022; Souto Ferreira et al., 2022; Matrajt et al., 2021b; Childs et al., 2022; Gonzalez-Parra, 2021; Tuite et al., 2021). One noteworthy ABM-based study uses differential vaccine function parameters for the Moderna and Pfizer-BioNTech vaccine, and finds that to minimize cases the second Moderna dose should be delayed while a delay of the second Pfizer-BioNTech dose may be detrimental if pre-existing immunity is low and if single dose-induced immunity wanes (Moghadas et al., 2021). To minimize deaths or hospitalizations, this study suggests delayed second doses, irrespective of the type of vaccine.

The various studies differ in how the two-dose vaccination campaign is implemented. In compartment-based models, separate compartments for singledose (*V*_1_) and “fully” vaccinated (*V*_2_) individuals are used. One common implementation employs two rates *ν*_1_(*t*)*, ν*_2_(*t*) *≥* 0 to describe the proportion of susceptible and single-dose vaccinated that receive a vaccine dose on a given day *t* (see e.g., Zhao et al. (2021b); Childs et al. (2022); Liu et al. (2022a)). That is,

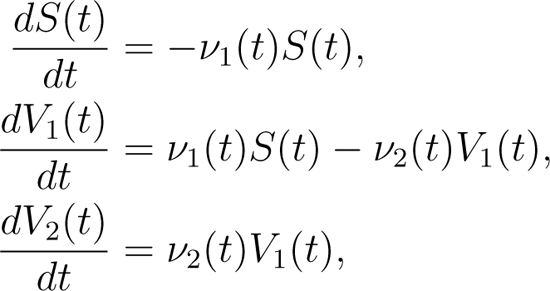

with constraints on *ν*_1_(*t*) and *ν*_2_(*t*) ensuring that only available vaccines are used. Other implementations include delay differential equations (Souto Ferreira et al., 2022; Sepulveda et al., 2023) or weekly pulse vaccinations (Matrajt et al., 2021b).

### 4.6. Transmission rates and heterogeneous contact patterns

The rate at which susceptible individuals acquire an infection, the force of infection, depends, among others, on contact rates, the community incidence and the infectivity of the virus. It is well-established that human interactions are age-assortative and that older individuals have on average fewer contacts (Mossong et al., 2008). A realistic account for age-specific mixing patterns is thus of paramount importance in infectious disease models that guide policy-makers to prioritize either high-contact young people or lower-contact older people. A common approach to model infection of sub-population *i, i* = 1*, …, n* in an age-structured ODE is

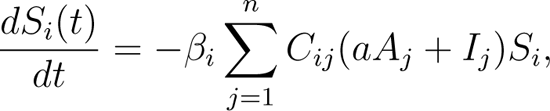

where

- *a ≥* 0 represents the relative contagiousness of asymptomatic (*A*) compared to symptomatic (*I*) individuals. All investigated studies chose *a ∈* (0, 1].
- The transmission rate *β_i_* can account for age-dependent susceptibility and risk mitigation (e.g., mask wearing). Multiple studies assumed that older, more vulnerable individuals suffer from higher susceptibility (Davies et al., 2020; Moore et al., 2021b; Jahn et al., 2021) but also engage in more risk mitigation measures (Masters et al., 2020; Kadelka and McCombs, 2021; Bushaj et al., 2023; Vo et al., 2023). The transmission rate may also vary over time, e.g., due to the emergence of more transmissible SARS-CoV-2 variants (Islam et al., 2021; Moore et al., 2021b), or time-varying social distancing levels (Moore et al., 2021b). It may further vary from location to location in spatially distributed models (Vo et al., 2023).
- The *n × n*-matrix *C* describes the average number of contacts an individual in sub-population *i* has with individuals from sub-population *j*. Just like *β_i_*, this matrix may also vary over time and by location to account for periods of school closures, work-from-home orders, etc. A reduction in activity levels of sub-population *i* (e.g., due to NPI adherence) can be implemented in two ways: (i) through a reduction in *β_i_*, or (ii) through a proportional reduction of row and column *i* of contact matrix *C*. It is very important to understand the differential effect of these choices on the model. Only the latter choice reduces both new infections of sub-population *i* and onward transmission by members of sub-population *i*. For this reason, this choice should be preferred in infectious disease models.

The seminal, diary-based POLYMOD study surveyed roughly one thousand individuals each in eight European countries and established country-specific contact matrices for a population stratified into 15 age groups (0 *−* 4, 5 *−* 9*, …,* 75 *−* 79, 80+) (Mossong et al., 2008). Contact rates were further stratified by location (home, workplace, school, other). By combining this data with various other data sources, age-and-location-specific synthetic contact rates were obtained for 177 countries, and it was shown that synthetic and empirical contact matrices employed in epidemiological models yield similar findings (Prem et al., 2017, 2021).

Most investigated mathematical models use a country-specific contact matrix. One ODE-based study shows explicitly how the optimal prioritization strategy depends on the country-specific age pyramid and contact matrix: to minimize deaths, it is optimal to first vaccinate the oldest people in India and Italy but middle-aged people in China (Wang et al., 2022). Another study quantifies the inter-generational mixing, which is typically lower in high-income countries (Gozzi et al., 2021). Since the population in high-income countries is on average also older, non-trivial dependencies arise when designing morbidity- or mortality-minimizing vaccine allocation strategies.

One commonly stated limitation of these contact matrices is that they have been derived before the COVID-19 pandemic, during which relative mixing patterns may have shifted. Two general approaches have been used to overcome this issue. First, several studies recomputed the overall contact matrix as a linear combination of the four location-specific pre-pandemic contact matrices (Matrajt et al., 2021b; Foy et al., 2021; Kiem et al., 2021; Chen et al., 2021; Moore et al., 2021b). That is,

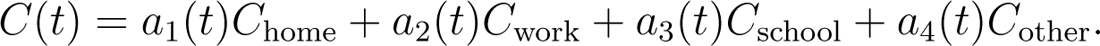

All studies agree that during a pandemic, *a*_1_(*t*) *≥* 1 while 0 *≤ a*_2_(*t*)*, a*_3_(*t*)*, a*_4_(*t*) *<* 1. Cell phone mobility data (Jentsch et al., 2021; Foy et al., 2021) as well as specific policy implementations (school closures, work-from-home orders, etc) (Jentsch et al., 2021; Karabay et al., 2021) have been used to inform the weights *a*_1_(*t*)*, …, a*_4_(*t*). The Oxford Stringency Indices are based on information on implemented government policies related to closure and containment, health and economic policy, and provide means to quantify the time-varying level of NPIs in 180 countries for several countries, even at the level of individual jurisdictions (Hale et al., 2021). Second, empirical contact matrices have been derived during the pandemic in several settings, and are used by a variety of studies, see e.g. (Han et al., 2021; Tran et al., 2021; Zhao et al., 2021b). Comparisons of Chinese as well as Belgian diary-based pre-pandemic contact matrices with contact matrices obtained during and after the first COVID-19 lockdowns revealed not only drastic changes in the number of contacts but also in the mixing patterns (Zhang et al., 2020, 2021; Coletti et al., 2020). Contact matrices during a pandemic specifically during periods of strong NPI adherence, e.g., a lockdown exhibit much lower levels of age-assortativity, likely due to school and work closures.

While age-assortativity decreases during a pandemic, assortative mixing with respect to other attributes, also known as homophily (McPherson et al., 2001), may be high or even increase, and may profoundly influence disease dynamics. High levels of homophily with respect to COVID-19 vaccine status have been reported (Are et al., 2024). Both network- and ODE-based studies show that this may lead, at a fixed level of vaccine coverage, to more frequent outbreaks and higher attack rates (Kadelka and McCombs, 2021; Hiraoka et al., 2022; Burgio et al., 2022). Another study, employing a novel approach to include homophily with respect to binary attributes in established contact matrices (Kadelka, 2023), shows that accounting for high levels of ethnic homophily in the United States, coupled with proportionately more people of color working in high-contact jobs but fewer being of old age, leads to non-trivial trade-offs in optimal vaccine prioritization design (Kadelka et al., 2022).

A less frequently mentioned limitation of contact matrices is that they are by default non-reciprocal. In empirical contact matrices, elderly tend to more frequently report a brief contact (Mossong et al., 2008), and generally provide less reliable responses in surveys Perry (1982). Physical contacts, which are required for COVID-19 spread, are however reciprocal. That is,

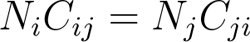

should hold for all *i, j* = 1*, …, n*, where *N_i_* is the size of sub-population *i*; otherwise, disease dynamics are inaccurate. A common procedure to generate a reciprocal contact matrix is outlined in (Funk et al., 2019; Kadelka, 2023). Several studies employ this or a similar procedure (see e.g. Matrajt et al. (2021b); Kadelka et al. (2022); Islam et al. (2021)).

### 4.7. Behavioral responses

The force of infection depends proportionately on the contact level. Over the course of a pandemic, implemented government policies, adherence to NPIs (e.g., social distancing), and thus contact levels differ. Variations in contact levels that affect the entire population homogeneously can be easily implemented by multiplying contact matrices with a time-varying factor, as described above. Heterogeneous behavioral responses, reported in several surveys (see e.g. Masters et al. (2020); Dryhurst et al. (2022); Pasion et al. (2020)), are often much harder to model but crucial to accurately predict, for example, the effect of a specific vaccine prioritization strategy. A variety of studies included aspects of homogeneous and heterogeneous behavioral responses.

Several studies assume that the population-wide contact level depends on the number of recent infections, hospitalizations and/or deaths (Rahmandad, 2022; Althobaity et al., 2022; Gozzi et al., 2021), the number of currently active cases (Jentsch et al., 2021; Islam et al., 2021), recent changes in these numbers, or a combination of these factors. For example in Islam et al. (2021), the population-wide contact level has been modeled to depend on the number of active cases, using a sigmoidal function. Another study, expanding the ODE model by Bubar et al (Bubar et al., 2021), assumes that the contact reduction depends exponentially on the number of deaths reported a few days ago (Rahmandad, 2022). The authors argue that inclusion of this endogenous behavioral feedback loop provides a better model fit to data. This study further assumes that the level of contact reduction can depend on age/perceived risk, and specifically, that vaccinated individuals may engage in less contact reduction. This complicates the trade-off in vaccine prioritization when minimizing deaths or YLL: vaccinate high-contact, less NPI-compliant individuals first or more compliant people at higher risk. This study concludes that the answer primarily depends on the speed of the vaccine roll-out, as well as the differences in NPI compliance. Several other studies come to the same or similar conclusion (Bushaj et al., 2023). An ABM-based study employs a binary stratification of NPI compliance, and quantifies how much lower the compliance level of low-risk (versus high-risk) individuals must be for them to be the optimal prioritization target (Kadelka and McCombs, 2021). It finds that the switching point depends on the vaccine efficacy, as well as the level of homophily with respect to vaccine status and NPI adherence. An ODE-based model, first proposed in Gozzi et al. (2021) and then adapted to several Arab countries in Althobaity et al. (2022), uses the same binary classification and explicitly models dynamic shifts between these two sub-populations that depend on the vaccine coverage and the number of recent deaths. These studies find that, at *R*_eff_ = 1.15, elderly should be prioritized when minimizing deaths as long as the vaccine roll-out is sufficiently fast; the speed at which the optimal prioritization switches is country-dependent. Some studies have been even shown that a vaccine with low effectiveness may be detrimental and yield worse outcomes than in the absence of a vaccine, due to behavior adaptation and a false belief of protection (Kadelka and McCombs, 2021; Luebben et al., 2023).

### 4.8. Vaccine hesitancy

Despite the availability of highly effective vaccines, a sizeable proportion of people refuses to get vaccinated against COVID-19. When determining the optimal roll-out of a limited vacccine, this factor must be taken into account, either explicitly by certain modeling assumptions or implicitly by only considering those roll-out solutions that appear feasible given levels of vaccine hesitancy. A number of survey-based studies have accessed these levels in different countries and at different times during the pandemic (Sallam, 2021; Soares et al., 2021). While vaccine hesitancy differs from country to country, it generally differs more with age. As expected, older individuals who are more at risk are also more willing to be vaccinated.

Many infectious disease models explicitly include vaccine hesitancy by assuming that a proportion of each sub-population cannot be vaccinated. Some models simply assume that this proportion is fixed (Bubar et al., 2021; Gavish and Katriel, 2022; Islam et al., 2021; Li et al., 2022; Luebben et al., 2023; Kadelka et al., 2022; Makhoul et al., 2020; Miura et al., 2021; Moore et al., 2021a; Rodriguez-Maroto et al., 2023; Tatapudi et al., 2021; Walker et al., 2022; Hogan et al., 2021; McBryde et al., 2021; Rahmandad, 2022), while others account for lower hesitancy among older individuals (Liu et al., 2022a; Moghadas et al., 2021; Han et al., 2021; Liu et al., 2022b; Moore et al., 2021b; Zavrakli et al., 2023). Most models in the latter category differentiate hesitancy using a binary age threshold. A Greek model infers age-specific values from a telephone survey (Sypsa et al., 2022; Barmpounakis et al., 2022). One study assumes that the level of hesitancy evolves over time following an ODE formulation (Jentsch et al., 2021). An ABM-based study shows that COVID-19 outbreaks are more frequent, at a given level of vaccine coverage and NPI adherence, if those who comply with NPIs are also those who get vaccinated, as is frequently the case (Kadelka and McCombs, 2021).

### 4.9. Effective reproductive number

While the basic reproductive number describes the number of secondary infections caused on average by an infected individual in a fully susceptible population, the effective reproductive number varies over time and takes into account population-wide levels of immunity, NPI adherence, emergence of more infectious variants, etc. Many models have investigated how optimal prioritization schemes depend on this key epidemiological parameter. As described above, prioritization of high-contact, low-risk younger individuals becomes a reasonable choice, when minimizing deaths, if the effective reproductive number is close to 1 (Chen et al., 2021; Althobaity et al., 2022; Gozzi et al., 2021; Bubar et al., 2021), especially if it is decreasing (Molla et al., 2022). A French study further clarifies that young individuals should only be prioritized if the vaccine acts almost entirely by reducing infections (that is, if *ɛ*_1_ *>> ɛ*_2_) (Kiem et al., 2021). In these circumstances, the vulnerable, older part of the population is indirectly protected by the vaccine through herd immunity, pending no changes in NPI adherence.

### 4.10. Variant Considerations

Like most RNA viruses, SARS-CoV-2 evolves rapidly (Markov et al., 2023). Over the course of the last four years, a plethora of variants has emerged. These variants exhibit different phenotypes characterized e.g. by transmissibility, severity of disease and immune evasion. The vaccine roll-out happened in parallel to the emergence of SARS-CoV-2 variants. For example, the United States started to vaccinate individuals in December 2020. By April 2021 when weekly vaccination counts still increased Alpha (B.1.1.7), which was an estimated 50% more transmissible, had become the dominating virus strain, followed soon after by Delta (B.1.617.2), which was even more contagious and caused also more hospitalizations and deaths (Campbell et al., 2021). Therefore, mathmematical models used to predict optimal vaccine allocation strategies should consider the emergence of variants. In practice, the time to emergence of a new variant and its specific phenotypic characteristics can, however, not be reliably predicted. Some studies used genomic SARS-CoV-2 surveillance time-series and estimates of the phenotypic characteristics of circulating variants to predict the future distribution of circulating virus strains (Islam et al., 2021; Childs et al., 2022). This distribution can yield estimates of the time-varying transmissibility and various transition probabilities, even when the model does not account explicitly for variant-specific infections (Liu et al., 2022a). A few studies went further and even included different compartments for those infected with e.g. the wildtype and Alpha variant (Aruffo et al., 2022; Gozzi et al., 2022). Qualitatively, accounting for the emergence of more infectious variants yields higher effective reproductive numbers (pending no changes in NPI adherence, etc) with effects as described in Subsection 4.9.

## 5. Summary of selected modeling studies

In the previous sections, we focused on key modeling assumptions, both explicit and implicit ones, and their impact on the outcomes of vaccine prioritization models. We highlighted different studies wherever suitable. In this section, we provide a brief summary of selected modeling studies that present interesting features. Only some features of the models and findings from the studies can be described. Subsection 5.1 summarizes studies that answer the question: How should a limited vaccine be optimally distributed among a population stratified by age? Subsection 5.2 summarizes studies that answer the question: Should the second COVID-19 vaccine dose be delayed given limited vaccine availability? Subsection 5.3 summarizes spatial vaccine distribution studies.

### 5.1. Summary of studies that employ an age-structured mathematical model

#### 5.1.1. Differential equation and difference equation based models

In Bubar et al. (2021), the authors compared five different vaccine prioritization strategies using an age-stratified ODE-based SEIR model. Outcomes were assessed using the number of infections, deaths and YLL. The authors found that prioritizing adults aged 20 to 49 years minimized infections at all considered values of the effective reproduction number (1.1-2). Furthermore, this same prioritization provided the best way to reduce mortality and YLL when the effective reproduction number is low (*≤* 1.15) and if the vaccine roll-out is slow. This study highlights the importance of the transmissibility of SARS-CoV-2 and the pace of vaccine roll-out on the choice of an optimal vaccination strategy. The authors also consider the potential benefit of seroprevalence tests prior to vaccination.

In Moore et al. (2021b), an ODE-based SEAIHR model was fitted to data from the United Kingdom. The authors show that even a vaccine as effective as those by Pfizer-BioNTech and Oxford-AstraZeneca would not suffice to contain the COVID-19 outbreak, partially due to age-varying vaccine hesitancy. The study furthers highlights that the number of deaths that appear among vaccinated will naturally increase as vaccine coverage increases. The model accounts for time-varying population-wide social distancing levels as well as the emergence of more transmissible variants.

In Buckner et al. (2021), the authors investigated optimal vaccination strategies by using an ODE-based SEPIAR model that takes into account essential workers. Stochastic nonlinear programming techniques were used to find the vaccine prioritization. Assuming *R*_eff_ = 2.5, vaccines were assigned only to susceptible individuals and updates to the prioritization were made each month. Outcomes were assessed using the number of infections, deaths and YLL. The authors found that to minimize infections, it is optimal to prioritize older essential workers. However, depending on the objective and alternative model scenarios considered, younger essential workers may be prioritized to control SARS-CoV-2 spread or elderly to directly control mortality. A combination of a genetic algorithm (global) and a simulated annealing algorithm (local) was used to obtain the optimal vaccination strategy each month.

In Foy et al. (2021), the authors employed an age-structured ODE-based SEIARQ (*Q* = quarantined, not spreading) model to inform the optimal vaccine roll-out in India. Assuming *R*_eff_ = 2.4, four vaccine prioritizations were compared: even across the population, prioritize 20–40 year olds, 40–60 year olds, or those 60 and older. To minimize deaths, the authors found that prioritizing the oldest is optimal regardless of the vaccine efficacy, control measures, vaccination pace, or immunity dynamics. However, this prioritization results in more symptomatic infections. To minimize infections, vaccination of 20-40 year olds should be prioritized. A faster vaccine roll-out reduces the differences between the compared vaccine prioritizations.

In MacIntyre et al. (2022), the authors employed an ODE-based SEPAIQR model that was extended to include several additional classes such as traced, undiagnosed and highly infectious. The model was fitted to the Australian state New South Wales and age-targeted and ring vaccination programs were compared. The population was stratified by occupation (healthcare workers) and age. The authors found that vaccinating older people prevents more deaths and that herd immunity can only be reached by mass vaccination campaigns, and only if the vaccine is sufficiently effective and rolled out sufficiently fast.

In Hogan et al. (2021), an extended SEIR discrete-time model was used to evaluate the public health impact of vaccines using data from different countries. The model uses a class of individuals with a mild infection that includes both symptomatic and asymptomatic but that does not require hospitalization. The authors identified death-minimizing vaccine allocation strategies withinand between-countries. They found if less than 20% vaccine coverage is available, it is better to prioritize the elderly. However, in less limited settings, high transmitters should be prioritized.

In Moore et al. (2021a), an ODE-based SEAIR model was used to investigate optimal vaccine allocations in the UK. Outcomes were assessed by deaths and loss in QALYs. For a range of model assumptions, the authors found elderly should be prioritized. However for vaccines that have low efficacy among the elderly (*<* 20%), other prioritizations proved more effective. In Shim (2021), an ODE-based SEIAR model was calibrated South Korea. The authors found that to minimize deaths (infections) older (younger) individuals should be prioritized. Interestingly, the YLL-minimizing strategy is sensitive to vaccine efficacy and the number of vaccine doses available. When vaccine efficacy (assuming a vaccine that only reduces infections) is relatively low (*≤* 30%) groups with high case-fatality rates should be prioritized, thereby maximizing the direct benefit of vaccines. However, with vaccines that have higher efficacy, prioritization shifts toward younger age groups: 40–69 year olds at 50–70% efficacy or 30–59 year olds at 90% efficacy.

In Islam et al. (2021), a detailed ODE-based model was calibrated to evaluate the U.S. vaccine roll-out. The population was stratified by age, comorbidity status, job type and living situation. The model also accounts for time-varying population-wide social distancing levels as well as the emergence of more transmissible variants. The authors compared 17.5 million 4-phased vaccine allocation strategies and found that a strategy that prioritizes people with comorbidities in all age groups is Pareto-optimal, yielding slightly fewer deaths, infections and YLL than the strategy recommended by the Centers for Disease Control (CDC).

In Penn and Donnelly (2023), an ODE-based SIR model was used to study the effect of the basic reproduction number *R*_0_ on the optimal vaccination plan. An interesting counter-intuitive result was found: It is better to prioritize 45–49 year olds than 55–59 year olds despite higher case fatality rates in the latter group. The authors explained this by the fact that the latter group has much fewer contacts with those 75 and older (as parents of those 55-59 years old have already died to a large degree). Thus, prioritization of 45–49 year olds substantially increases the secondary protection of the elderly. This result shows the importance of the age-stratified contact matrices.

In Zhao et al. (2021b), three different ODE-based SEIAR models were used to find the optimal vaccination strategy against COVID-19 in Wuhan City, China. The authors used the effective reproduction number to estimate the SARS-CoV-2 transmission between age groups. They found that, before NPIs were implemented, the highest transmissibility existed among those 15-44 years old. In order to control transmission, this age group should be prioritized. To minimize deaths, those *≥* 65 years old should be prioritized, irrespective of their lower contact rates.

In Matrajt et al. (2021a), an ODE-based model with many compartments was used to determine which age group(s) should be vaccinated assuming instantaneous vaccination and 10-100% vaccine coverage. The authors studied many scenarios and found that for low vaccine effectiveness (10-50%), regardless of vaccination coverage, it is optimal to prioritize elderly people when minimizing deaths. However, for higher vaccine effectiveness and if the basic reproductive number is low, it is better to prioritize younger people, especially if available vaccination coverage is *≥* 40%. The optimization routine includes a coarse global search algorithm, coupled with a fast optimizer, to explore the entire space of possible combinations of vaccine allocations.

In Kadelka et al. (2022), an age-and-ethnicity-stratified ODE-based model was used to study the optimal distribution of available vaccines in the United States to two different groups: White and Asian persons and all others. Different levels of ethnic homophily were considered. The authors found that vaccine allocations that stratify vaccine access by ethnicity could have prevented a number of deaths, especially assuming high levels of ethnic homophily. Moreover, the authors highlight a second trade-off when designing mortality-minimizing vaccination plans and accounting for ethnic homophily: the elderly population is predominantly White and Asian, while those employed in high-contact occupations are predominantly from the other ethnic groups.

In Stafford et al. (2023), an age-and-race-stratified ODE-based model was used to study the distribution of available vaccines in the United States to two different groups: non-Hispanic White persons and all others. Several objective functions that include mortality, YLLs, measures of inequity and joint disease burden were considered. The authors found that there exists a trade-off between minimizing disease burden and minimizing inequity, especially if vaccine is very limited (e.g., 10%). If vaccine coverage is *≥* 30%, both inequity and mortality can be optimized at the same time.

In Zuo et al. (2022), an ODE-based SEIQR model was used, combined with google mobility data to modify contact matrices. This study highlights that the optimal vaccine prioritization depends on particular parameters related to the transmission rates. Assuming fixed daily doses, the authors found that in a scenario with low infection rate and low vaccine availability, vaccinating first people over 60 minimizes deaths, but that with more vaccine availability vaccinating first those 51-60 year old is preferable due to their higher contacts.

In Gavish and Katriel (2022), an ODE-based model is used to investigate whether children should have been vaccinated earlier. The authors found that prioritization strategies that include vaccination of children lead to Paretooptimal outcomes regarding minimizing deaths and infections, especially if the basic reproductive number is high.

In Rao and Brandeau (2021b), an ODE-based SIR model with two age groups (with age threshold 65) was used to study which vaccine allocation minimizes the effective reproduction number. Assuming that all vaccinations take place at once, the authors found that the answer depends on available vaccine coverage, vaccination pace and the initial effective reproduction number. In Rao and Brandeau (2021a), the same authors used the model to minimize infections, deaths, YLL and loss in QALY. They found that it is better to prioritize the young group to minimize infections, but the older individuals for all other metrics. This result was obtained by simple analytical conditions that describe the optimal vaccine allocation for each objective.

In Rahmandad (2022), an ODE-based SEIR model was used to study the effects of behavioral responses to risk by means of an endogenous feedback loop. Specifically, the author assumed that population-wide social distancing levels fluctuate depending on the recently reported numbers of COVID-19 deaths. The author argues that high-contact individuals should be prioritized to minimize deaths or YLLs, as long as the vaccine roll-out happens fast enough. This is because the vulnerable population is already more risk-averse and thus engages in more risk mitigation.

In Han et al. (2021), an ODE-based SIR model was used to study optimal vaccine prioritization plans in China. The authors show that a time-varying vaccination program (i.e., allocating vaccines to different target groups as the epidemic evolves) can yield much better outcomes since it is capable to simultaneously achieve different objectives (e.g., minimizing deaths and infections). In addition, a high vaccination pace in the early phase of the vaccination plan is better. In a sensitivity analysis, the authors employed a contact matrix derived from contact diaries collected in Shanghai in March 2020, at a time when the lockdown was over, but severe NPIs were still in place. The “pandemic” contact matrix exhibits much less age-assortativity.

In Makhoul et al. (2020), an ODE-based model was used and the authors found that a vaccine with efficacy against infection *≥* 70% would eliminate COVID-19. Outcomes were assessed over the course of ten years and the authors assumed full vaccine protection over this time course, which appears too high retrospectively. The authors studied two vaccination programs: 80% coverage before the onset of the epidemic and 80% coverage within one month of the onset of the epidemic.

In Campos et al. (2021), an ODE-based SIR model was used to predict the COVID-19 dynamics and compare with out-of-sample data from Rio de Janeiro. In addition, numerical simulations were used to compare age-based vaccine allocation strategies policies. Three age groups of similar size were considered as vaccination targets. In in all the tested scenarios, prioritization should be given to either those 15-34 or 50 year and older. The optimal choice depends on the evaluation time period, vaccination schedules and efficacy of the vaccine.

In Angelov et al. (2023), a non-standard age-structured ODE-based model was proposed that differentiates between isolated and non-isolated as well as symptomatically and asymptomatically infected. The model further takes into account the heterogeneity of the infected sub-population with respect to the time since infection. Solving an optimal control problem, which considers as one of the constraints the hospital capacity, the authors found that deaths in Austria are minimized if those 18-30 years old (highest transmitters) are vaccinated first, followed by those 80 and older (most at risk), followed by other age groups.

In Babus et al. (2023), a linear programming problem is solved in order to find a U.S. vaccination plan that minimizes deaths and the economic cost of a stay-at-home order. The study considered occupation-based risk exposure (454 occupations). The authors compared three different plans. Under the only considered plan without a stay-at-home order, the largest number of vaccines should be allocated to those 50–59 years old, followed by those 60–69. In general, the best plans focused on age-based risk rather than occupation-based risk exposure.

In Miura et al. (2021), the age-specific transmission intensities (i.e., the next generation matrix) are reconstructed using an approximation method. This enables the inference of the expected impact of vaccinating each subgroup from data on incidence and force of infection. This unique approach requires only routine surveillance data on the number of cases to determine the best possible allocation of vaccines, and can be employed in data-scarce environments. The method is tested with data from the Netherlands. The authors conclude that the optimal timing of changing from vaccinating one age group to another depends on the specific objective.

In Cartocci et al. (2021) an ODE-based SIR model that considers timevarying parameters and sex was used to compare Italian vaccination programs, using the outcomes YLL, deaths and infections. According to the model, deaths (infections) are minimized by prioritizing elderly (younger). However, the optimal YLL-minimizing strategy depends on the effective reproductive number. If it is high, younger individuals should be prioritized.

In Galli et al. (2023) an ODE-based SIR model was used to predict COVID-19 dynamics and evaluate vaccination plans in the Southwest Shewa Zone in Ethiopia. A plan that prioritizes those 50 years and older was found to avoid more critical cases than a random vaccine allocation.

In Gonźalez-Parra et al. (2022), two ODE-based SIAR models were used to study vaccine allocation strategies. Different scenarios related to the speed of the vaccine roll-out were compared. The authors found that generally those 55 years and older should be prioritized to minimize deaths. However, whenever the transmission rate is relatively high and elderly have a substantially lower transmission rate than younger people, the optimal prioritization switches.

In Aruffo et al. (2022), an ODE-based model with many compartments is used to study different Canadian vaccine roll-out and NPI-lifting scenarios. To minimize infections and shorten the time until NPIs can be lifted, those 20-59 years old should be prioritized. Different reopening scenarios and strategies were compared, with total cases and deaths depending on the timing of lifting NPIs.

#### 5.1.2. Agent-based models (ABMs)

ABMs offer more flexibility and potential realism than ODE-based models but a proper analysis of these stochastic models requires simulations and is thus computationally expensive.

In Jahn et al. (2021), an ABM was developed to derive optimal vaccine allocation strategies for Austria. The model contains 9 million agents, one for each Austrian resident. Each agent possesses an associated state variable that describes its disease state. The model further accounts for age, occupation (health care workers), testing and notification delays. The probability of an infection occurring during a single contact between an infected and a susceptible was determined by calibrating the model to the number of detected Austrian COVID-19 cases in March 2020. The authors found that hospitalization and deaths were minimized if elderly and vulnerable were prioritized, assuming very limited vaccine availability. To assign more vaccines, the authors highlight the usefulness of a stepwise optimal allocation technique, in which small batches of vaccine are assigned at a time.

In Ben-Zuk et al. (2022), an ABM was used to derive and compare optimal vaccine allocation strategies for two Israeli cities of similar size but with different household size and age distributions. The authors compared two strategies: vaccinate those prioritized by public health decision makers, or dynamically prioritize neighborhoods with a high estimated reproductive number. Using infections and deaths as outcomes, the study highlights that optimal vaccination plans depend on subpopulation-specific infection rates and unique demographic characteristics.

In Kadelka and McCombs (2021), an ABM was used to highlight the effect of homophily and correlation between attitudes and opinions on vaccine prioritization. The authors argue that the U.S. society exhibits high levels of homophily w.r.t. to vaccine willingness and NPI adherence and that these two attributes are correlated, i.e., that people who get vaccinated are also more likely to engage in other risk mitigation. The authors found that these attributes must be taken into account to inform the optimal vaccine prioritization strategy, as they influence at which relative contact level of older compared to younger individuals the optimal prioritization target switches. In Tatapudi et al. (2021), an ABM that considers various NPIs was developed to track the number of COVID-19 cases, hospitalized, and deaths for all age groups. 2.8 million agents were used to represent each resident in Miami Dade County, United States. Three vaccine allocation strategies were compared: (i) random allocation, (ii) prioritization by age, (iii) a minor variant of the CDC strategy, which prioritizes health care workers in addition to elderly. The authors found that a random allocation minimizes infections, while the CDC strategy minimizes deaths and YLL, although it proved only slightly better than the other two strategies.

In Bushaj et al. (2023), the Covasim ABM from Kerr et al. (2021) was expanded to compare a random with an age-structured vaccine allocation strategy. The authors show that a “governor Deep Reinforcement Learning agent” can learn effective strategies and suggest, based on a multi-objective reward structure, optimal ABM interventions when presented with a specific epidemic situation. Moreover, the study shows that focused vaccination of super-spreaders can substantially reduce infections at the expense of marginally more total deaths. The model was tested with data from the U.S. states New Jersey and Kansas.

In Cattaneo et al. (2022), the Covasim model is used to determine the number of infections and deaths prevented by vaccines in the Italian region Lombardy, and to retrospectively evaluate vaccine allocation strategies. Prioritization of the elderly and at-risk categories, as used in Italy, was validated as the most effective in reducing deaths, however only as long as the vaccine roll-out happens fast enough.

### 5.2. Summary of optimal COVID-19 vaccine dosing interval studies that employ a mathematical model

The following studies all use a mathematical model that differentiates between those vaccinated with a single dose and two doses (i.e., fully vaccinated). The fundamental vaccine prioritization trade-off is between vaccinating more people at lower levels of protection or inducing higher protection for fewer individuals.

In Moghadas et al. (2021), an age-structured ABM with compartments SEPIAR was used that differentiated between the Pfizer and the Moderna vaccine. Varying rates of vaccine-induced immunity waning were considered. In addition, maximum vaccine coverage (i.e., vaccine hesitancy) was assumed to be age-dependent. Model parameters were informed by data from the United States. Outcomes were assessed by infections, hospitalizations, and deaths. The authors found that a delay of the second dose of at least 9 weeks would have averted deaths and hospitalizations compared to the recommended 4-week interval. For infections, the results differed for the two considered vaccines: while a delay of the second dose of Moderna vaccines would have reduced infections, delaying second doses of Pfizer vaccines may have caused more infections if pre-existing immunity is below 30% and if vaccine-induce one-dose immunity wanes.

In Tuite et al. (2021), a decision analytic cohort model was used to assess strategies for dose allocation (assuming a steady vaccine supply). The authors found that variants of a flexible strategy that keeps only 10% of the supply for second doses during the first 3 weeks are better than the fixed strategy employed by the United States.

In Souto Ferreira et al. (2022), an age-structured SEAIHR delay differential equation model was used to study the optimal timing between first and second dose. A constant vaccine production rate was assumed and vaccination rates were optimized using linear programming, with outcomes assessed by deaths. The authors found that the best strategy depends on an interplay between the vaccine production rate and the relative efficacy of the first dose.

In Ferreira et al. (2022), a discrete-time compartmental model, fitted to Brazil and differentiating between three different vaccines, was used to investigate the optimal vaccine priortization and dosing interval, which was varied from 8 to 12 weeks. The authors found that a shorter time interval between first and second dose for the AstraZeneca vaccine would minimize deaths. However, in their analysis, it appears that the vaccine availability is not fixed, i.e., a shorter time interval corresponds to more available vaccine, which is obviously beneficial. Moreover, the authors assumed large differences in vaccine efficacy between the first and the second doses, contrary to many other studies.

In Zuo et al. (2022), an ODE-based SEIQR model was fitted to South Africa and used to answer questions related to vaccine priortization and delay of the second dose. The authors found that, assuming limited vaccine availability, a delay of second doses leads to fewer severe COVID-19 cases.

In Gianatti et al. (2023), a discrete-time model with compartments SEPIHR (no age groups) is fitted to data from the city of Tandil, Argentina. Assuming constant numbers of daily available vaccines, different fixed delays between the vaccine doses (28, 42, 72 days) were compared using death as the outcome metric. An optimal control problem was solved to determine the best way to administrate the available vaccines, by considering two controls that represent the number of first and second doses applied each day. The authors found that delaying the second dose as long as possible (72 days in the study) was optimal.

In Mak et al. (2022), an ODE-based SEPAIHR model was used to investigate three different policies related to vaccine roll-out: holding back second doses, releasing second doses, and delaying the time between doses. The authors found that releasing second doses reduces infections. However, stretching the between-dose time flattens the infection curve and reduces both hospitalizations and mortality compared with a strategy that releases second doses. The model includes details related to the inventory dynamics of the vaccine roll-out process not found in other models. The authors further conduct extensive sensitivity analyses related to age composition, risk-based prioritization, supply disruptions, and disease transmissibility.

In Romero-Brufau et al. (2021), an age-structured ABM was used to investigate the effect of a delayed second dose on deaths, infections and hospitalizations. A total of 100k agents interact in 3 types of networks (occupation, family and random) over a period of six months. In all compared vaccination plans, the allocation started with the oldest group and proceeded by decreasing age. The authors found that a delayed second dose yields lower deaths as long as the first dose is sufficiently effective (*≥* 80%) but that a delay does not affect the YLL and infections much.

In Diarra et al. (2022), an ODE-based SEIARQ model, an adaptation of the CoMo model (Aguas et al., 2020), was used to study vaccination strategies in Senegal. Three particular vaccination strategies were evaluated, using deaths as outcome metric. The authors found the second dose should be delayed for those 40 years or younger.

In Childs et al. (2022), an age-structured ODE-based SEIS model that considers reinfections and immunity was used to determine questions related to vaccine prioritization and delay of the second dose in Canada. The authors found that a delay, as well as earlier vaccination of 15-19 year olds would both yield lower infections numbers.

### 5.3. Summary of spatial vaccine distribution studies that employ a mathematical model

In Grauer et al. (2020), a computational model with Brownian agents moving randomly through a continuous square space with periodic boundary conditions was introduced. Each agent has an internal state variable describing its disease state (e.g., S, I, or R). A statistical mean-field model was applied to study three vaccine allocation strategies: (i) distribution of vaccines proportional to population density, (ii) an “infection weighted strategy” that distributes vaccines proportional to the quantitative value of the bi-linear incidence rate *βSI*, and (iii) a “focusing strategy” that distributes the vaccines sequentially by prioritizing the regions with the highest incidence rate. The authors found that the last strategy minimized deaths; age was however not considered.

In Molla et al. (2022), a spatial ODE-based model was developed to model COVID-19 disease dynamics in five different Finnish regions. The authors combined age-specific contact data with geographic movement data to investigate the optimal vaccination strategies. Using optimal control methods, the authors found that allocating vaccines demographically and in an age-descending order is not optimal for minimizing deaths or infection cases. Instead, it proved optimal to prioritize high-incidence regions and allocate vaccines at the same time to different age groups.

In Zhou et al. (2021), the authors used cell phone data from a Chinese city to develop a spatial ABM for a realistic urban scenario. To compare seven different scenarios related to vaccine allocation, the authors assigned the vaccines by fulfilling the priority group before advancing to the next priority group. The authors found that the vaccine coverage to reach herd immunity varies strongly across locations, highlighting the immense usefulness of knowledge of the spatial heterogeneity when designing vaccine allocation strategies.

In Lemaitre et al. (2022), an ODE-based spatial model of the 107 Italian provinces, originally developed in Gatto et al. (2020), was used to study optimal vaccine distribution across space. Google Community Mobility Reports was used to estimate the variations in mobility across provinces and as a proxy for changes in social contacts. The authors developed a novel optimal control framework that yields the best vaccination strategy under realistic supply and logistics constraints. The identified optimal strategy, which substantially outperforms standard strategies, has a complex structure: while mainly dependent on the projected incidence of each province, it also takes into account the spatial connectivity between provinces.

In Vo et al. (2023), an age-stratified ODE-based spatial SEIR model of the 50 U.S. states was used to illustrate the utility of mechanistic expressions for the basic and effective reproductive number, as well as to compare two vaccine prioritization strategies: a uniform allocation and an allocation along the gradient of the effective reproductive number. The authors showed that the latter approach yields fewer infections but they acknowledged that this would come at the expense of more hospitalizations and deaths.

## 6. Related studies that employ optimal control methods

The majority of studies included in this review identified vaccine allocation strategies that optimize a given metric, e.g. minimizing deaths or infections. Some studies went further and identified strategies that are Pareto-optimal with respect to multiple objectives, see e.g. Islam et al. (2021); Gavish and Katriel (2022); Diarra et al. (2022). A number of studies, some already described above, went even further and employed classical optimal control theory to find vaccination strategies that minimize a variety of health and/or economic outcomes. Some of these studies even consider age-dependent vaccine access.

These studies define a functional often a linear combination of different metrics that is optimized given some constraints, e.g., to account for limited vaccine availability. A general challenge of optimal control approaches is the high sensitivity of the resulting optimal vaccination strategy on the choice of weights in the functional. Moreover, the choice of functional itself affects the results. Nevertheless, these studies can provide important insights as the setup is more flexible, and we briefly describe some interesting approaches and note that several others (e.g., Lemaitre et al. (2022); Angelov et al. (2023)) are already summarized above.

In Acuña-Zegarra et al. (2021), an ODE-based SEAIR model (no age structure) was used to show that the optimal vaccination strategy depends on the speed of the vaccine roll-out and the length of natural immunity. The transmission contact rates and proportion of symptomatic cases were estimated by calibrating the model to observed death counts. The basic reproductive number was estimated to be in the range of [3.30, 4.84]. The authors minimized a functional that was a linear combination of YLL and Years Lost due to Disability. The authors found that varying the number of doses during the vaccine roll-out (if supply allows) yields to better outcomes than an approach with fixed number of vaccinations per day.

In Tu et al. (2023), the authors proposed a reaction-diffusion COVID-19 model (no age structure) to investigate how different vaccination-isolation strategies impact the COVID-19 pandemic. The functional included three metrics: social cost, social benefit, and the basic reproduction number. The authors found that for a given social cost or benefit, there are many Pareto-optimal vaccination-isolation strategies. The proposed model considered also a spatial variable, in addition to parameters related to social distancing and vaccination.

In Olivares and Staffetti (2021a), two control variables, vaccination and testing, were used to find the optimal strategy that minimizes a functional that accounts for the number of infected people with life-threatening symptoms and the number of deaths. The underlying model is ODE-based with a variety of compartments. Several optimal control problems were solved for different scenarios. Among others, the authors found that it is optimal to roll-out a vaccine as fast as possible. In Olivares and Staffetti (2021b), the same authors studied scenarios with different vaccine availability. The functional here depends on the number of symptomatic and asymptomatic infectious. The authors found again that early implementation of vaccination and testing reduces the number of symptomatically infected the most. However, if vaccine availability increases gradually, the optimal vaccination strategy differs quite strongly from other scenarios. Finally in Olivares and Staffetti (2021c), the same authors considered a mass vaccination plan, and polynomial chaos expansion was used to assess the uncertainty of the modeling outcomes.

In Ziarelli et al. (2023), an age-stratified two-dose ODE-based SIR model was calibrated to death counts from Italy, and several optimal control problems were solved, minimizing deaths, infections and hospitalizations independently. In each problem, the total number of vaccine doses was fixed but the distribution of the available doses among susceptibles and those who already received their first dose was optimized. The authors found that the deaths-minimizing strategy prioritized those 80 years and older, followed, interestingly, by those 20-39 years old. On the other hand, the infections-minimizing vaccination strategy prioritizes the 20-39 and 40-59 age groups but not children and teenagers despite them having the most contacts. This work nicely highlights the complexities of designing optimal age-based vaccine prioritization strategies.

In Choi and Shim (2021), an age-structured ODE-based model for South Korea was developed. Solving an optimal control problem with a functional that considers the cost of vaccination, as well as the cost of symptomatic and hospitalized infected, the authors found that the optimal vaccination strategy depends on the way the vaccine functions. While “susceptibility-reducing” vaccines should be allocated relatively evenly. On the other hand, “symptom-reducing” vaccines should, surprisingly, be allocated to those 20-29 and 50 and older but not to those 30-49 years old. The impact of vaccine function proved particular strong if the roll-out was assumed to be fast.

In Libotte et al. (2020), an SIR model was calibrated to data from China. An inverse problem was solved to determine the transmission rate, infectious period, initial number of infecteds and basic reproduction number (*R*_0_). The authors developed a multi-objective optimal control problem, in which the number of vaccines and the total number of infected are simultanenously minimized. This problem is solved using Differential Evolution, yielding a set of Pareto-optimal vaccination strategies.

In Zhang et al. (2024), an optimal control problem was solved with the aim of minimizing deaths and conserving vaccines at the same time. The population was divided into four subpopulations: health workers, young individuals, middle-aged individuals, and the elderly. The authors found that the optimal vaccination strategy substantially improved upon a proportional vaccine roll-out.

There exist numerous other works that use classical optimal control to identify optimal COVID-19 vaccination strategies, most of them minimizing an objective functional which accounts for infected cases, deaths or the number of vaccines Agossou et al. (2021); Al-arydah (2023); Salcedo-Varela et al. (2023); Shen et al. (2021); Zaitri et al. (2022). In particular, some works have combined optimal control with age-structured models to find the optimal vaccination allocation Avcı and Yurtŏglu (2023); Chhetri et al. (2022); Kumar et al. (2021). Optimal control employed on infectious disease models represents a powerful tool to identify optimal vaccine allocation strategies. However, setting up the optimal control problem including the constraints regarding vaccine availability is crucial but it is challenging to restrict the search to vaccination programs that can be implemented in the real world. The choice of the functional to be minimized is also crucial, as strongly affects the optimal outcomes, see e.g., Ledzewicz and Schättler (2020).

## 7. Conclusion

The COVID-19 pandemic constitutes one of the worst pandemics humankind has ever endured, both in terms of lives lost and economic repercussions. It is also the first pandemic in a globalized world. The rapid spread of the disease around the world was enabled by high levels of connection, transport and travel between distant parts of the world. This is not going to change, which is why the world will eventually face another pandemic. Whether this will be caused by a highly transmissible SARS-CoV-2 that has evolved to evade immune defenses or by an entirely novel pathogen cannot be predicted. However, we can learn from mistakes made during the COVID-19 pandemic to ensure better preparedness for a future pandemic. On the mathematical modeling front, this includes fully understanding the effect realistic human behavior and social processes have on the outcomes in infectious disease models. Specifically for models designed to inform prioritization strategies for a vaccine that will initially always be limited, we need to look beyond the details of specific models and understand the greater connections behind explicit and implicit model assumptions and outcomes. This is what we attempted in this systematic review of mathematical models designed to find optimal COVID-19 vaccine prioritization strategies.

## Data Availability

All data produced in the present work are contained in the manuscript

## Acknowledgments

G.G.-P. acknowledges funding from Maria Zambrano (UPV, Ministry of Universities of Spain by the European Union-Next generation EU) and an Institutional Development Award (IDeA) from the National Institute of General Medical Sciences of the National Institutes of Health under grant number P20GM103451. C.K. was partially supported by a travel grant (712537) from the Simons Foundation.

